# Development of second and third-trimester population-specific machine learning pregnancy dating model (Garbhini-GA2) derived from the GARBH-Ini cohort in north India

**DOI:** 10.1101/2021.10.02.21264450

**Authors:** Nikhita Damaraju, Ashley Xavier, Ramya Vijayram, Bapu Koundinya Desiraju, Sumit Misra, Ashok Khurana, Nitya Wadhwa, GARBH-Ini Study Group, Raghunathan Rengaswamy, Ramachandran Thiruvengadam, Shinjini Bhatnagar, Himanshu Sinha

## Abstract

**Background:** The prevalence of preterm birth (PTB) is high in lower and middle-income countries (LMIC) such as India. In LMIC, since a large proportion seeks antenatal care for the first time beyond 14-weeks of pregnancy, accurate estimation of gestational age (GA) using measures derived from ultrasonography scans in the second and third trimesters is of paramount importance. Different models have been developed globally to estimate GA, and currently, LMIC uses Hadlock’s formula derived from data based on a North American cohort. This study aimed to develop a population-specific model using data from GARBH-Ini, a multidimensional and ongoing pregnancy cohort established in a district hospital in North India for studying PTB.

**Methods:** Data obtained by longitudinal ultrasonography across all trimesters of pregnancy was used to develop and validate GA models for second and third trimesters. The first trimester GA estimated by ultrasonography was considered the Gold Standard. The second and third trimester GA model named, Garbhini-GA2 is a multivariate random forest model using five ultrasonographic parameters routinely measured during this period. Garbhini-GA2 model was compared to Hadlock and INTERGROWTH-21st models in the TEST set by estimating root-mean-squared error, bias and PTB rate.

**Findings:** Garbhini-GA2 reduced the GA estimation error by 23-45% compared to the published models. Furthermore, the PTB rate estimated using Garbhini-GA2 was more accurate when compared to published formulae that overestimated the rate by 1·5-2·0 times.

**Interpretation:** The Garbhini-GA2 model developed is the first of its kind developed solely using Indian population data. The higher accuracy of GA estimation by Garbhini-GA2 emphasises the need to apply population-specific GA formulae to improve antenatal care and better PTB rate estimates.

**Funding:** Centre for Integrative Biology and Systems Medicine, IIT Madras; Department of Biotechnology, Government of India; Grand Challenges India, BIRAC.

**Panel: Research in Context:** *Evidence before this study:* The appropriate delivery of antenatal care and accurate delivery date estimation is heavily dependent on accurate pregnancy dating. Unlike GA estimation using crown-rump length in the first trimester, dating using foetal biometry during the second and third trimesters is prone to inaccuracies. This is a public health concern, particularly in low and middle-income countries like India, where nearly 40% of pregnant women seek their first antenatal care beyond 14 weeks of gestation. The dating formulae used in LMIC were developed using foetal biometry data from the Caucasian population, and these formulae are prone to be erroneous when used in ethnically different populations.

*Added value of this study:* This study developed a dating model, the Garbhini-GA2 model for second and third trimesters of pregnancy using multiple candidate biometric predictors measured in a North Indian population. When evaluated internally, this model outperformed the currently used dating models by reducing the errors in the estimation of gestational age by 25-40%. Further, Garbhini-GA2 estimated a PTB rate similar to that estimated by the Gold Standard in our population, while the published formulae overestimated the PTB rates.

*Implications of all the available evidence:* Our Garbhini-GA2 model, after due validations in independent cohorts across the Southeast Asian regions, has the potential to be quickly translated for clinical use across the region. A precise dating will benefit obstetricians and neonatologists to plan antenatal and neonatal care more exactly. From an epidemiologist standpoint, using the Garbhini-GA2 dating formulae will improve the precision of the estimates of pregnancy outcomes that heavily depend on gestational age, such as preterm birth, small for gestational age and stillbirth in our population. Additionally, our dating models will improve phenotyping by reducing the risk of misclassification between outcomes for mechanistic and biomarker research.

## Introduction

Estimation of gestational age, commonly known as pregnancy dating, is the cornerstone of obstetric care. Delivery of antenatal care – either investigation for foetal morphological anomalies, gestational diabetes, and preeclampsia, or nutritional supplementation and foetal growth monitoring is primarily dependent on accurate dating. From an epidemiological standpoint, dating of pregnancy determines the accuracy of population-level estimates of pregnancy outcomes such as preterm birth, foetal growth restriction, and stillbirth. Conventionally, counting the number of days from the last menstrual period (LMP) using Naegle’s methods (1) has been used to arrive at the estimated delivery date and, thereby, gestational age. This method is still used as the primary method in many low and middle-income countries. The use of LMP to compute GA is highly contingent on factors such as regularity of menstrual cycle and accurate recall of the date of LMP. The menstrual cycle’s regularity is susceptible to change in conditions like polycystic ovarian syndrome (2) and obesity (3). Consumption of contraceptives and breastfeeding just before conception can also affect the menstrual cycle (4,5).

The best time and method for assessing GA during pregnancy is the first trimester using ultrasonographic measurement of crown-rump length (CRL) (1,6–9). The measurement protocols for CRL are well established and practised as a standard of care globally. The physiological and pathological variations in foetal growth can affect the accuracy of GA prediction in the second and third trimesters (10). GA dating formulae based on foetal biometry in the second and third trimesters, such as Hadlock’s formula (11), are widely used in USG machines in India. However, Hadlock’s formulae were developed on a sample of pregnant Caucasian women from a North American population. Foetal growth can have ethnic variations in the South Asian population, where foetuses are generally smaller than their western counterparts. This may lead to underestimating GA in our setting by models developed from a western population (12). This underestimation may cause delays in planning deliveries and consequently an increased risk of post-dated pregnancy, perinatal asphyxia, and stillbirth.

The ideal way of developing and overcoming the influence of foetal growth in dating is to identify biometric features that are relatively less affected by foetal growth restriction. An accurate dating model for the second and third trimesters is essential in LMICs like India, where more than 40% of pregnant women seek antenatal care beyond their first trimester (13).

In this study, we developed dating models for second and third trimesters of pregnancy using multiple candidate biometric predictors measured in a North Indian population and validated it internally in a separate dataset from the same population. We evaluated the impact of the newly developed model on the estimation of preterm birth (PTB) rate and compared it against globally published models.

## Methods

### Study design and data collection

The GARBH–Ini cohort (interdisciplinary Group for Advanced Research on BirtH outcomes-DBT India Initiative) is an ongoing prospective observational cohort of pregnant women initiated in May 2015 at Gurugram Civil Hospital, Haryana, India. The participants were enrolled before 20 weeks of gestation and followed three times during pregnancy (18-20w, 26-28w and 30-32w) till delivery. Detailed methods of the GARBH-Ini study have been published earlier (14). A first trimester dating scan was performed if a participant was enrolled within 14 weeks of her pregnancy. She was followed at least once in each trimester till the end of the pregnancy, when an ultrasound examination was performed to measure foetal biometry and other foetal and maternal characteristics. Participants in the cohort were enrolled after obtaining written informed consent. Ethics approvals were obtained from the institutional ethics committees of Gurugram Civil Hospital, Safdarjung Hospital, Translational Health Science and Technology Institute, and Indian Institute of Technology Madras (14).

The dataset for this study was derived from the GARBH-Ini study consisting of 6498 participants at the time of enrolment. We included participants who had their pregnancy dating done in their first trimester, had undergone one ultrasonographic examination in the second or third trimester of pregnancy and had the outcome of their pregnancy documented (n = 2649). The details of selection criteria and participant flow are provided in Figure 1. Data collected for a participant in the second and third trimesters were treated as different observations leading to a sample of N_o_ = 4972. Of these observations, 4768 observations were selected based on the availability of all five ultrasound metrics – Biparietal diameter (BPD), occipitofrontal diameter (OFD), head perimeter (HP), abdominal perimeter (AP) and femur length (FL).

**Figure 1:**
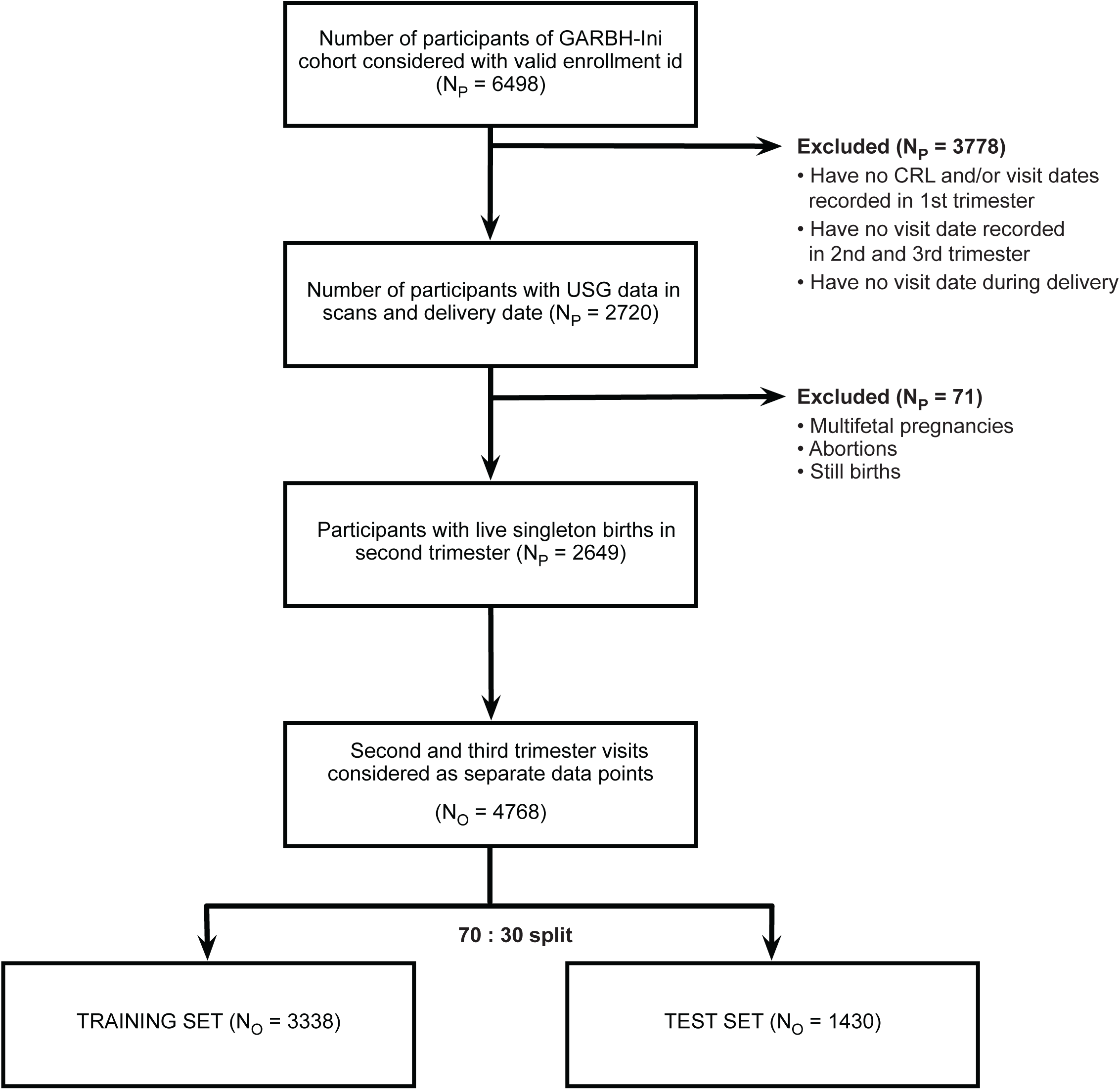
Outline of the data selection process for TRAINING and TEST set. Exclusion criteria for each step are indicated. N_p_ indicates the number of participants included or excluded by that criterion, and N_o_ shows the number of unique observations derived from the participants in a dataset.

### Dataset preparation for modelling and internal validation

The resultant dataset consisting of 4768 observations was split into a TRAINING set of 3338 observations (70% of the dataset and an unseen TEST set of 1430 observations (30% of the dataset, see Figure 1). The unseen TEST set was used for internal validation to compare the performance of our models with published ones.

### Definition of gold standard GA

Gestational age for participants at their second and third-trimester visits was computed using the GA estimated during their first trimester dating scan. In our previous work (15), we had developed a CRL based first-trimester dating model (Garbhini-GA1) and showed it to be performing as well as the Hadlock’s and INTERGROWTH-21st first trimester dating models. We used the Garbhini-GA1 model to obtain the first trimester dating for our study participants. The difference in the visit dates of the first and second trimester scans was added to the first trimester GA (estimated by Garbhini-GA1) to give the second and third trimester GA used for all subsequent analyses (Figure S1). These GA estimates were considered the Gold Standard or ground truth for modelling our second and third-trimester formulae.

### Feature selection

Feature selection was implemented on a set of 21 candidate features (Table S1), including the five main USG variables (BPD, OFD, HP, AP & FL). Boruta, a wrapper built on a random forest-based classifier using the boruta package (16), was used to implement feature selection. Features selected by Boruta undergo a robust method of computing relative importance, a metric that indicates the usefulness of a feature in predicting GA. In each iteration, a feature is compared with a randomly shuffled version of itself called a shadow feature (Supplementary Methods). The highest feature importance recorded among all randomly shuffled shadow features called shadowMax was used as a threshold to decide if a feature was selected (Figure 2A).

**Figure 2:**
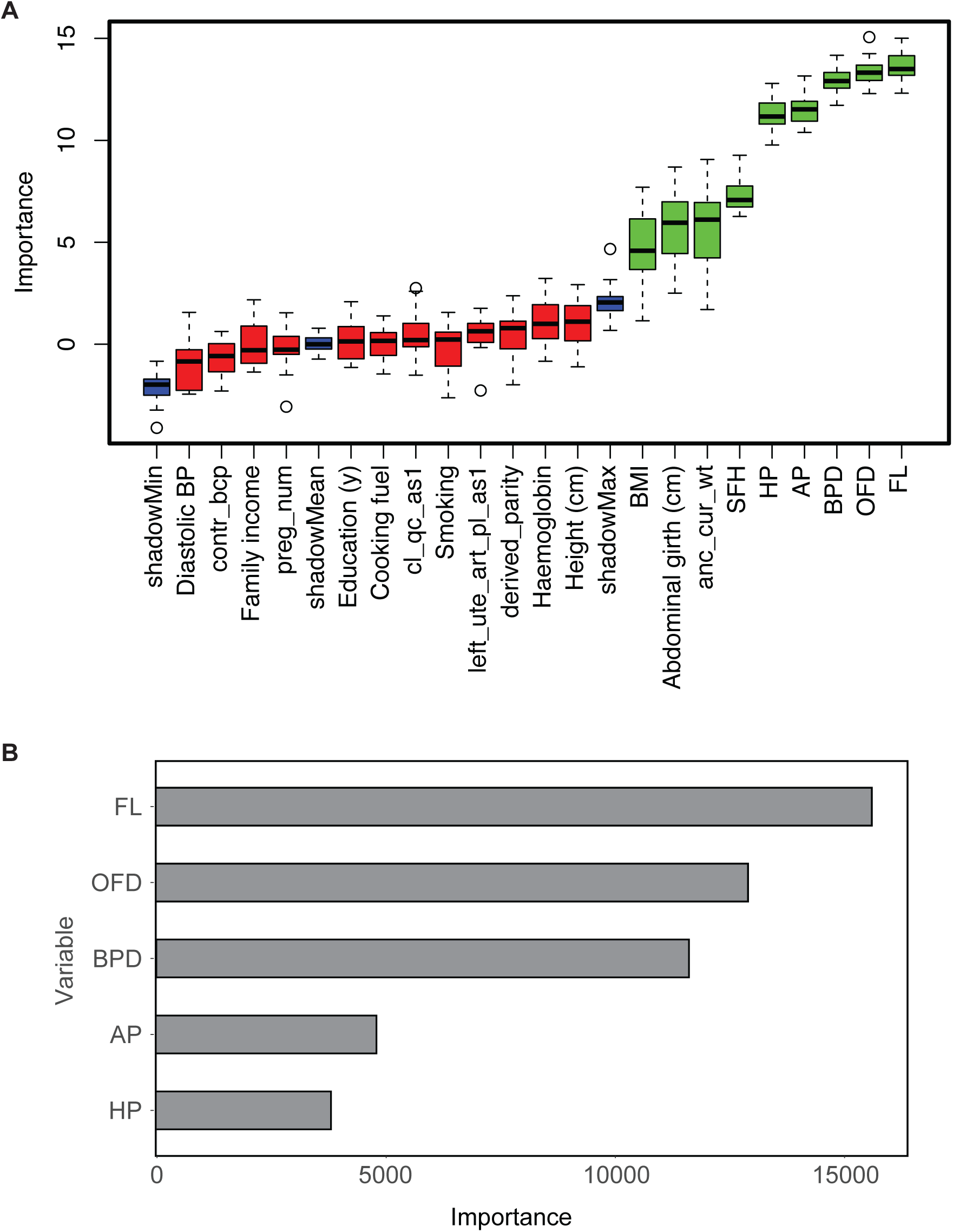
Feature selection and ranking of their importance. (A) Feature selection: Variable importance graph from the output of Boruta shows the relative importance with a 95% confidence interval of importance estimate for each of the 22 features in predicting GA. Green box plots correspond to the features that are selected while red box plots correspond to those that are rejected. Blue box plots represent the minimum, average and maximum value for a shadow attribute used to select features. (B) Variables in order of importance in the Garbhini-GA2 Random Forest model: A bar graph denoting important variables as measured by recording the decrease MSE each time a variable is used as a node split in a tree in the Random Forest model. A variable with higher importance will have the largest average decrease in MSE across all the 500 trees. The variables are denoted on the y-axis, and the relative importance of the variable is denoted in the x-axis.

### Development of population-specific gestational dating model

We used three different methods to develop a second and third trimester dating model for our dataset – polynomial regression, random forest, and gradient boosting machines.

Random forest was implemented using the ranger package (17). The number of trees was decided by observing the number of trees contributing to the least mean squared error (MSE). Hyperparameters mtry (number of variables to randomly sample as candidates at each split), minimum node size (minimum number of samples within terminal nodes), sample fraction (proportion of the dataset to train in each iteration) were tuned using a grid search that computes the out-of-bag (OOB) RMSE value for 140 sets of parameters. The optimal model was then used to identify the relative importance of features in decreasing order (Figure 2B).

Polynomial model search, and Gradient Boosting methods were also implemented as potential GA estimating models (see Supplementary Methods).

### Comparison of dating models

The best fit models from polynomial model search, Random Forest and Gradient Boosting developed on the TRAINING set were compared with Hadlock’s best performing formula (11) consisting of BPD, FL, HP and AP (GA = 10·85 + 0·060(HP)(FL) + 0·67(BPD) + 0·168(AP)), and INTERGROWTH-21st (18) formula consisting of HP and FL (log_e_(GA) = 0·03243 × (log_e_(HP))^2^ + 0·001644 × FL × log_e_(HP) + 3·813). The performance of each model was calculated by computing the root-mean-square deviation RMSE on the TEST set (N_o_ = 1430) by calculating the mean of residual (difference between predicted model and gold standard) for each data point and taking the square root of that mean. Parity plots for each model were constructed by plotting the predicted GA on the y-axis and Gold Standard GA on the x-axis. Violin plots constructed using the ggplot2 package (19) were used to demonstrate the distribution of errors of estimation of GA (predicted GA – actual GA) for all the models. The model with the least RMSE, based on random forest modelling, named the Garbhini-GA2 model, was picked for further comparative analyses. Using Bland-Altman analysis, the bias between different formulae was evaluated, and pairwise mean difference and limits of agreement were reported.

A series of datasets comprising 1000 simulated data points were generated to understand a model’s predictions visually. Each dataset was constructed by generating random values for a particular USG parameter while keeping the other four a constant at the mean of their value in the TEST set. This process was repeated to generate simulated datasets specific to the second and third trimesters as well. Three models – Garbhini-GA2, Hadlock and INTERGROWTH-21st were applied onto the simulated datasets to compare predicted GA versus USG variables visually.

Using predicted GA from Garbhini-GA2 and Hadlock’s formula, two groups of participants were created from the TEST set – having an absolute difference in predicted GA of less than 1-week and greater than 1-week, respectively. Descriptive statistics for continuous and categorical variables (see Table 1) were generated to compare the two groups.

**Table 1:** Baseline characteristics of the participants included in the TRAINING (N_p_ = 1803), and TEST (N_p_ = 772) sets.

| **Sociodemographic characteristics** | **TRAINING set**<br>**Median (IQR) or N (%)**<br>**or Mean ± SD** | **TEST set**<br>**Median (IQR) or N (%)**<br>**or Mean ± SD** |
| --- | --- | --- |
| Age (years) | 23 (21,26) | 23 (21,26) |
| GA at enrolment by USG (weeks) | 20.9 ± 4.2 | 20.9 ± 4.2 |
| BMI at enrolment into the cohort |  |  |
| Underweight | 27.8 % | 24.4 % |
| Normal | 57.2 % | 61.0 % |
| Obese | 12.0 % | 11.0 % |
| Overweight | 1.9 % | 2.3 % |
| Haemoglobin (g/dL) | 9 (8.5 - 9.5) | 9 (8.4 - 9.5) |
| Weight (kgs) | 56.5 (51.0 - 63.4) | 57.1 (51.5 - 63.4) |
| Height (cm) | 153.1 (149.4 - 157) | 152.7 (149.2 - 156.5) |
| Socioeconomic Status |  |  |
| 0 | 0.4 % | 0.6 % |
| 1 | 18.1 % | 19.3 % |
| 2 | 37.9 % | 33.4 % |
| 3 | 42.8 % | 45.6 % |
| 4 | 0.2 % | 0.5 % |
| Undetermined | 0.5 % | 0.5 % |
| Parity (number) |  |  |
| 0 | 51.0 % | 50.2 % |
| 1 | 33·6 % | 32·4 % |
| 2 | 12·1 % | 13·5 % |
| 3 | 2·6 % | 3·2 % |
| 4 | 0·7 % | 0·6 % |
| 5 | 0·1 % | 0 |
| 6 | 0 | 0 |
| 7 | 0 | 0 |
| Level of education |  |  |
| Illiterate | 18·2 % | 17·8 % |
| Literate or primary school | 9·6 % | 12·7 % |
| Middle school | 16·0 % | 13·9 % |
| High school | 22·5 % | 22·4 % |
| Post high school diploma | 17·1 % | 17·5 % |
| Graduate | 13·1 % | 13·5 % |
| Post-graduate | 2·9 % | 1·8 % |
| Occupation |  |  |
| Unemployed | 92·8 % | 91·4 % |
| Unskilled worker | 3·2 % | 4·8 % |
| Semi-skilled worker | 1·4 % | 1·7 % |
| Skilled worker | 1·9 % | 1·2 % |
| Clerk, shop, farm owner | 0·1 % | 0·1 % |
| Semi-professional | 0·3 % | 0·1 % |
| Professional | 0·2 % | 0·6 % |
| Religion |  |  |
| Hindu | 91·6 % | 91·4 % |
| Muslim | 7·2 % | 6·7 % |
| Sikh | 0·4 % | 0·5 % |
| Christian | 0·7 % | 0·9 % |
| Buddhist | 0·0 % | 0·2 % |
| More than one religion | 0·1 % | 0·1 % |
| Other | 0·0 % | 0·0 % |
| Fuel used for cooking |  |  |
| Biomass fuel | 93·0 % | 92·5 % |
| Clean fuel | 7·0 % | 7·5 % |
| Source of drinking water |  |  |
| Safe water | 56·6 % | 58·8 % |
| Unsafe water | 43·4 % | 41·2 % |
| Second-hand tobacco smoke |  |  |
| Exposed | 19·4 % | 18·5 % |
| Unexposed | 0·1 % | 81·4 % |
| Undetermined | 80·5 % | 0·1 % |
| History of any chronic illnesses |  |  |
| Absent | 98·3 % | 97·5 % |
| Present | 1·7 % | 2·5 % |
| History of hypertensive disease of pregnancy |  |  |
| Absent | 98·9 % | 99·1 % |
| Present | 1·1 % | 0·9 % |
| History of contraceptive at the time of conception |  |  |
| Absent | 95·9 % | 95·1 % |
| Present | 4·1 % | 4·9 % |

### Preterm birth analyses

For classification of PTB, the number of participants that had a GA less than 37 weeks per 100 participants was calculated and tabulated for the three formulae, namely Hadlock, INTERGROWTH-21st, Garbhini-GA2 and Gold-Standard GA on the TEST set. Further, the 95% confidence interval for each estimate was computed using the Clopper-Pearson method using the binom package (20). The agreement between the Gold Standard and three formulae for preterm labelling was calculated using the Jaccard similarity coefficient. Classification of participants as preterm was graphically depicted using a quadrant plot consisting of predicted GA on the x-axis and gold standard GA on the y-axis divided by a line at 37 weeks, respectively. All modelling and statistical analysis were performed using the R programming language.

## Results

### Description of participants included in the study

The median age of the participants enrolled in the study was 23 years in the TRAINING and TEST sets. The median weight and height in the TRAINING set was 56·5 kg (IQR 51·0 - 63·4), and 153·1 cm (IQR 149·4 - 157), respectively and 57·2 % of participants had a normal BMI (median 20·4 IQR 18·2 - 23·1). More than half of them were primigravida (51·0 %). Most of the participants were from the middle or lower socioeconomic strata (21). The participants selected in this study had a median GA of 19·4 weeks (IQR 19·1 - 20·1). Other baseline characteristics are given in Table 1.

### Feature selection

Feature selection by Boruta assigned importance to all the 22 features used in the analysis and selected nine that crossed the threshold (shown in green in Figure 2A), namely - BPD, OFD, FL, HP, AP, symphysiofundal height, BMI, maternal weight, and abdominal girth. The remaining features were rejected (Figure 2A) as they did not cross the threshold level of importance. Further, the relative importance of all features showed that the five USG based metrics - AP, FL, OFD, HP, and BPD had a marked higher significance than non-USG based metrics. USG-based metrics were used to develop GA models in this study.

### Order of importance of features

The variable importance in the random forest model was recorded by computing the decrease in MSE each time a variable was used to split a tree at a node. Variables that reduce the remaining error the most after a node is split were considered more important than others. The overall importance was computed by averaging the decrease in MSE across all trees in the forest. Most impactful variables were considered to be of most importance (Figure 2B).

### Comparison of published methods and Garbhini formulae in the second and third trimesters

Comparisons between different dating models showed that the root-mean-square deviation (RMSE) on the TEST set varied between 0·89 - 1·57 weeks. Our Garbhini-GA2 model had the least error (0·89 weeks, 95% CI: 0·80, 0·97 weeks) and Hadlock’s formula the maximum of 1·57 weeks with the INTERGROWTH-21st model in between at 1·16 weeks (Figure 3A, Table 2). This demonstrated that the error with the Garbhini-GA2 model was substantially lower than the Hadlock’s (by 45%) and INTERGROWTH-21st (by 23%) models. Pairwise BA analysis between all formulae ranged between -0·642 and 1·089, with the largest difference between Hadlock’s formula and Garbhini-GA2 (Table 3). The Hadlock formula underestimated GA by a week as compared to the Garbhini-GA2 model. The INTERGROWTH-21st model too underestimated GA, albeit with a lesser magnitude (3 days).

**Table 2:** Performance characteristics of the GA estimation models compared to the Gold Standard GA on TEST set. An extended table with a comparison of other models developed is given in Table S2.

| **Model** | **RMSE with 95% CI** |
| --- | --- |
| Hadlock | 1·57 [1·52, 1·60] |
| INTERGROWTH-21st | 1·16 [1·08, 1·23] |
| Garbhini-GA2 | 0·89 [0·80, 0·97] |

RMSE is the root mean squared error of GA estimated by each model compared to Gold Standard GA in weeks.

**Table 3:** Bland Altman analysis and PTB agreement on TEST set. Each value in the bottom diagonal consists of a pairwise mean difference between the formulae with each other and with respect to the Gold Standard GA. The value in brackets represents the limits of agreement. Each value in the top diagonal is the agreement between the Gold Standard and three formulae for preterm labelling computed using the Jaccard similarity coefficient. An extended table with a comparison of other models developed is given in Table S3.

|  | **Gold Standard** | **Hadlock** | **INTERGROWTH-21st** | **Garbhini-GA2** |
| --- | --- | --- | --- | --- |
| **Gold Standard** |  | 41·486 | 60·280 | 78·750 |
| **Hadlock** | 1·089 (-1·125, 3·302) |  | 64·596 | 46·894 |
| **INTERGROWTH-21st** | 0·447 (-1·66, 2·554) | -0·642 (-1·599, 0·315) |  | 72·596 |
| **Garbhini-GA2** | 0·027 (-1·718, 1·772) | 1·061 (-0·251, 2·374) | 0·42 (-0·702, 1·541) |  |

**Figure 3:**
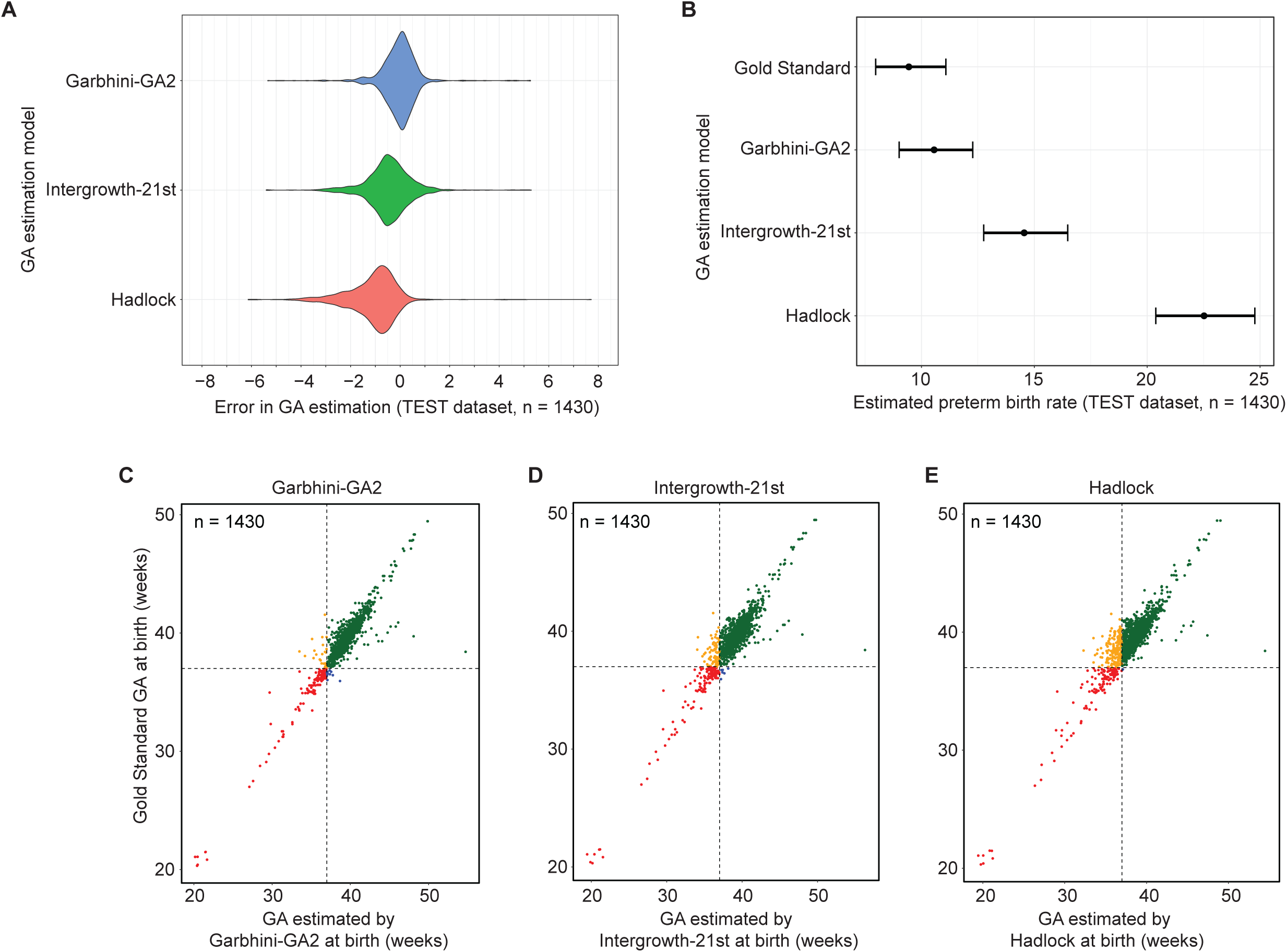
Performance of Garbhini-GA2 model in TEST set. (A) Distribution of error (in weeks) in the estimation of GA compared to the Gold Standard GA. The x-axis is the error (the difference between predicted and Gold Standard GA), the y-axis is the density for each of the models in the TEST set (N_o_ = 1430). (Extended figure to be added in Supplementary section). (B) PTB rates by various models: PTB rates labelled by each model with 95% confidence intervals on TEST set (N_o_ = 1430). (C-E) Comparison of individual-level classification of preterm birth by a model and Gold Standard GA. Green (term birth for both), red (preterm birth for both), blue (term birth for Gold Standard but preterm birth for model) and purple (term for model but preterm for Gold Standard).

We evaluated the contribution of foetal biometry towards the discrepancy observed among the three models using simulated datasets generated for the second, third and both second and third trimesters (see Methods). Our analyses showed that while differences were observed for the second and third trimester, it was slightly more for the second trimester (Figure S6). All the foetal biometry features seem to have contributed to the difference. Furthermore, the foetal biometry features were significantly lower for participants with an estimated GA difference of more than 1-week compared to those with less than 1-week difference (see Table S8).

### Impact of choice of dating formula on the estimation of preterm rates

The PTB rate in the TEST set using the gold standard for GA was 9·4% (CI 8·0, 11·1). The PTB rates estimated when different models did the dating of pregnancy ranged between 10·6 and 22·5% when computed on the TEST set. The PTB rates in the TEST set, when Garbhini-GA2 (10·6%; CI 9·1, 12·3) and Garbhini-XG Boost (10·6%, CI 9·0, 12·3) were used, were closest to those estimated by the Gold Standard (first trimester CRL-based) dating method. Remarkably, INTERGROWTH-21st (14·5%; CI 12·8, 16·5) and Hadlock’s formula (22·5%; CI 20·4, 24·8) overestimated PTB rates (Figure 3B, Table S4). The improved accuracy of Garbhini-GA2 was confirmed even when we redefined the Gold Standard using the Hadlock and INTERGROWTH-21st first trimester formulae (Table S7).

In terms of which births were classified as PTB, the Garbhini-GA2 dating model showed the highest (78·8%), and Hadlock’s formula showed the lowest agreement (41·5%) with the Gold Standard dating method (Table 3). This indicated that the use of Garbhini-GA2 resulted in the least number of misclassified births compared to the published formulae (Figure 3C).

## Discussion

In this study, we successfully developed the Garbhini-GA2 model, using routine foetal biometry such as BPD, OFD, HC, AP and FL to estimate GA in the second and third trimesters of pregnancy with an accuracy closest to the first trimester CRL-based dating. The Garbhini-GA2 model demonstrated 40% less error than the commonly used Hadlock’s formula and nearly 25% less than the most recent INTERGROWTH-21st formula. We show our Garbhini-GA2 formula is a better fit for the foetal biometry of our population. The implication of this improved accuracy of GA assessment was remarkably evident when we estimated PTB rates. The Hadlock’s and INTERGROWTH-21st formulae overestimated the PTB rate in our TEST population. However, the PTB rate estimated by the Garbhini-GA2 model was the closest to that estimated using the Gold Standard CRL-based dating model. This was due to a lesser degree of misclassification between preterm and term birth by Garbhini-GA2.

In India, the enrolment rate for antenatal care in the first trimester is low, with nearly 40% of the pregnant women getting their dating done in the second or third trimesters (13). The need for an accurate dating model for second and third trimesters, in this scenario, has been well recognised. Dating by LMP is inaccurate in the later trimesters due to poor recall, and that by ultrasonography is mired with an underestimation due to wide variations in foetal growth. The two approaches used to address this challenge are – first, using biometric parameters that may be relatively less influenced by variations in foetal growth, such as trans-cerebellar diameter (22) and second, developing models using a study population that has representation from LMIC (18). While the first approach is desirable, novel biometric measurements come with the challenges of measurement standardisation and retraining of sonologists, thereby delaying their translation. We used the second approach because it uses routine and well-standardised biometric measurements, which are quickly and widely translatable to clinical practice.

Among the three models we evaluated (Garbhini-GA2, Hadlock and INTERGROWTH-21st), it was not surprising that the Hadlock formula had the maximum error and bias. Hadlock formula was constructed using data from a low-risk North American Caucasian population. The variation in foetal biometry attributed to ethnic differences between North American and Indian populations was probably well captured in our Garbhini-GA2 model, improving its accuracy. The INTERGROWTH-21st formula was developed using representation from multi-ethnic populations, including those from India and other Southeast Asian nations and therefore performed better than the Hadlock formula but still was less accurate than Garbhini-GA2. The Garbhini-GA2 model had the least error, in comparison to Hadlock or INTERGROWTH-21st late trimester formulae, in estimating PTB rates regardless of whether the Gold Standard (CRL-based) dating was derived using Garbhini-GA1, Hadlock or INTERGROWTH-21st first trimester formulae. This least error showed that Garbhini-GA2 had the highest accuracy in estimating PTB rates. Among the five biometric parameters, FL, BPD, and OFD were the most important features contributing to the Garbhini-GA2 model. The head (BPD and OFD) and femur measurements were probably more homogeneous in our population than the abdominal circumference, and the former measurements change as a function of gestational age.

While our validation exercise in an unseen dataset kept aside during the model-building exercise showed improved performance of the Garbhini-GA2 model, it is well known that models perform optimistically when tested in a dataset derived from the same population. As we intend our models to be used across India and probably extended to the Southeast Asian region, we will evaluate them in multiple other populations. Another limitation of our work is that most of the data we had are from 18-22 weeks in the second and 30-32 and 35-37 weeks in the third trimester. We aim to overcome this limitation during the validation phases.

Garbhini-GA2 model with improved accuracy is primarily intended to be used by clinicians for dating pregnancy in women who seek antenatal care for the first time beyond 14 weeks of gestation. We believe, if validated externally, this will be an important intervention in assuring accurate dating and thereby help to time the delivery better. Precise dating will also benefit neonatologists to plan neonatal care, particularly for preterm neonates. From an epidemiologist standpoint, using the Garbhini-GA1 and Garbhini-GA2 dating formulae will improve the precision of the estimates of pregnancy outcomes that heavily depend on gestational age, such as preterm birth, small for gestational age and stillbirth in our population. Beyond clinicians and epidemiologists, our population-specific dating models will benefit biologists by improving the clinical phenotyping of these birth outcomes for mechanistic and biomarker studies.

## Conclusion

A late trimester dating model that can provide GA estimation as accurate as first-trimester dating will be a significant intervention to improve birth outcomes. Our Garbhini-GA2 model almost halved the error in the estimation of GA in the second and third trimesters compared to the commonly used Hadlock model. After due validations in independent cohorts across the Southeast Asian regions, our model has the potential to be quickly translated for clinical use across the region.

## Data Availability

The datasets used and analysed in the current study are available from the corresponding author upon reasonable request. All the codes used for this paper are available at https://github.com/HimanshuLab/Garbhini-GA2.

## Acknowledgement

We thank all the participants of the GARBH-Ini study. We thank members of the Centre for Integrative Biology and Systems Medicine (IBSE), Robert Bosch Centre for Data Science and Artificial Intelligence (RBCDSAI), IIT Madras, and Aryabhata Data science and AI Program at THSTI (ADAPT), THSTI. We also thank Balaraman Ravindran, IIT Madras, Gagandeep Kang, Christian Medical College, Vellore, India, and Ashok Venkitaraman, National University of Singapore, Singapore, for their valuable suggestions.

Members of GARBH-Ini Study Group: Translational Health Science and Technology Institute, NCR Biotech Cluster, Faridabad, India-Coordinating Institute (Vineeta Bal, Shinjini Bhatnagar (PI), Bhabatosh Das, Mahadev Dash, Bapu Koundinya Desiraju, Pallavi Kshetrapal, Sumit Misra, Uma Chandra Mouli Natchu, Satyajit Rath, Kanika Sachdeva, Dharmendra Sharma, Amanpreet Singh, Shailaja Sopory, Ramachandran Thiruvengadam, Nitya Wadhwa); National Institute of Biomedical Genomics, Kalyani, West Bengal, India (Arindam Maitra, Partha P Majumder (Co-PI) Souvik Mukherjee); Regional Centre for Biotechnology, NCR Biotech Cluster, Faridabad, India (Tushar K Maiti); Clinical Development Services Agency, Translational Health Science and Technology Institute, NCR-Biotech Cluster, Faridabad, India (Monika Bahl, Shubra Bansal); Gurugram Civil Hospital, Haryana, India (Umesh Mehta, Sunita Sharma, Brahmdeep Sindhu); Safdarjung Hospital, New Delhi, India (Sugandha Arya, Rekha Bharti, Harish Chellani, Pratima Mittal); Maulana Azad Medical College, New Delhi, India (Anju Garg, Siddharth Ramji), The Ultrasound Lab, Defence Colony, New Delhi, India (Ashok Khurana); Hamdard Institute of Medical Sciences and Research, Jamia Hamdard University, New Delhi, India (Reva Tripathi); All India Institute of Medical Sciences, New Delhi, India (Alpesh Goyal, Yashdeep Gupta, Smriti Hari, Nikhil Tandon); Government of Haryana, India (Rakesh Gupta); International Centre For Genetic Engineering and Biotechnology, New Delhi, India (Dinakar M Salunke Co-PI); G Balakrish Nair (Rajiv Gandhi Centre for Biotechnology, Trivandrum); Gagandeep Kang (Christian Medical College, Vellore).

## Supplementary material

Supplementary material document is available for this manuscript.

