## Supplementary Material for "Development of second and third-trimester population-specific machine learning pregnancy dating model (Garbhini-GA2) derived from the GARBH-Ini cohort in north India"

### Table of Contents

| | |
|---|---|
| Methods | 2 |
| Supplementary figure legends | 3 |
| Supplementary figures | 4-7 |
| Supplementary tables | 8 |

**Methods**

*Polynomial Regression and Gradient Boosting*

In addition to Random Forest, two methods - Polynomial regression and Gradient boosting were implemented. Polynomial regression was implemented using a brute force approach by taking all combinations in the form of logarithmic, square root, linear, quadratic, cubic and fourth power polynomial for the five main USG features – BPD, OFD, HP, FL and AP. For each formula, 100 iterations of bootstrapping were performed. In each iteration, 70% of the TRAINING set was randomly sampled to train the formula. The remaining 30% of the TRAINING set was used to fit the formula and compute the Adjusted $R^2$, $R^2$ and RMSE values (Figure S2). For each formula, the average of all the three metrics was added to a table if the formula was seen in more than 80% of iterations.

Gradient boosting was implemented using the xgboost package. The optimal number of trees was decided by minimising the loss function of interest coupled with cross-validation. Several sets of hyperparameters, namely shrinkage (controls the rate of descent), interaction depth (number of splits in each tree) and bag fraction (proportion of the dataset to be used to train in iteration), were tuned using a grid search method. The optimal model was selected by picking the parameters with the lowest five-fold cross-validated error on the TRAINING set. This model was used to identify the relative importance of features in decreasing order (Figure S3A). The gradient boosting algorithm computes the improvement in MSE for regression and averages the improvement across all the trees used to compute the relative importance. The variables that show the largest overall decrease in MSE have the highest importance. The preterm birth rate was estimated as 10.6% [9.1,12.3]. Classification of participants as preterm was graphically depicted using a quadrant plot consisting of predicted GA on the x-axis and gold standard GA on the y-axis divided by a line at 37 weeks (Figure S3B).

*Post-term birth analyses*

For post-term births, the number of participants with a GA of more than 42 weeks per 100 participants was calculated and tabulated for the three formulae, namely Hadlock, INTERGROWTH-21st, Garbhini-GA2 and gold-standard GA on the TEST set. Further, the 95% confidence interval for each estimate was computed using the Clopper-Pearson method using the binom package (*BinomCI Function | R Documentation*, n.d.). The agreement between the gold standard and three formulae for post-term labelling was computed using the Jaccard similarity coefficient. Classification of participants as post-term was graphically depicted using a quadrant plot consisting of predicted GA on the x-axis and gold standard GA on the y-axis divided by a line at 42 weeks. Post-term birth rates in our TEST set as gold standard dating method was 4.3% (95%CI: 3.3, 5.4). Post-term birth rates estimated by different models ranged between 2.7 and 5.8% (Table 6), with Hadlock's formula estimating the least (2.7%; CI 1.9, 3.7), followed by Intergrowth (4.5%, CI 3.5, 5.7), Garbhini-XG Boost (5.7; CI 4.6, 7.1) and Garbhini-GA2 being the highest (5.8%; CI 4.6, 7.1) (Table S5).

As with preterm birth, the Gold Standard shows maximum agreement with post-term birth classification done by Garbhini-GA2 and least with Hadlock's formula (Table S6). This indicates that using the Garbhini-GA2 model results in the least number of misclassified post-term births compared to the Gold Standard (Figure S4).

*Impact of choice of gold standard on the error in estimation and PTB rates*

Comparisons between different dating models when alternate gold standards were used showed that the root-mean-square deviation (RMSE) on the TEST set ranged between 0.77 - 1.24 when Hadlock's first-trimester dating formulae (22) was used and 0.75 - 1.21 when the INTERGROWTH-21st first trimester dating formula (23) was used. In both cases, Garbhini-GA2 was observed to have the least RMSE and Hadlock's formula with the highest RMSE. This indicates that the Random Forest approach is most accurate in estimating GA with alternate gold standards (Table S7 and Figure S5).

PTB rates estimated by different models on the TEST set ranged between 13.0 and 23.9%, 11.5 and 21.5% for Gold Standards as Hadlock's formula and INTERGROWTH-21st, respectively (Figure S5). The highest Jaccard similarity coefficient with respect to the Gold Standard in both scenarios was observed for Garbhini-XG Boost (0.72 and 0.74 for Hadlock's and Intergrowth formula, respectively,) followed closely by Garbhini-GA2 (0.71 and 0.73). The least Jaccard coefficient was observed for Hadlock's formula in both scenarios (0.52 and 0.52).

**Supplementary Figure Legends**

**Figure S1** Flowchart showing calculation of Gold Standard GA for each observation

**Figure S2** Flowchart showing methodology for the development of linear, quadratic, cubic and degree 4 polynomial models. Each model consists of all combinations of 5 USG features - BPD, OFD, HP, AP and FL, resulting in 4096 formulae.

**Figure S3** (A) Variables in decreasing order of importance by gradient boosting model. Variables in order of importance in the Garbhini-XG Boost model: To compute the relative importance, the gradient boosting algorithm computes the improvement in MSE for regression and averages the improvement across all the trees used. The variables that show the largest overall decrease in MSE have the highest importance. The variables are denoted on the y-axis, and the relative importance of the variable is denoted in the x-axis.
(B) Comparison of individual-level classification of preterm birth by XG Boost and gold standard GA. Green (term birth for both), red (preterm birth for both), blue (term birth for gold standard but preterm birth for model) and purple (term for model but preterm for gold standard).

**Figure S4** Comparison of individual-level classification of post-term birth by a model and Gold Standard GA. Green (term birth for both), red (post-term birth for both), blue (term birth for gold standard but post-term birth for model) and purple (term for model but post-term for gold standard).

**Figure S5** PTB rates calculated by different models using the Gold Standard as (A) Hadlock's first trimester formula (B) INTERGROWTH-21st first trimester formula on the TEST set

**Figure S6** Simulated data analysis. Each figure shows the prediction of three models – Garbhini-GA2, Hadlock's formula and INTERGROWTH-21st on the three sets of simulated datasets – (A, B) Second and third trimester combined (C, D) Second trimester (E, F) Third trimester

### Figure S1

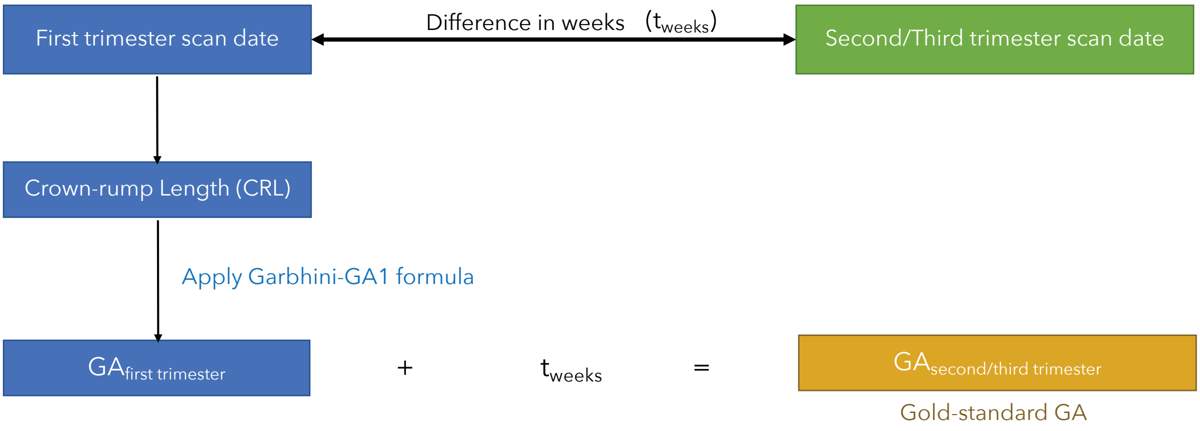


### Figure S2

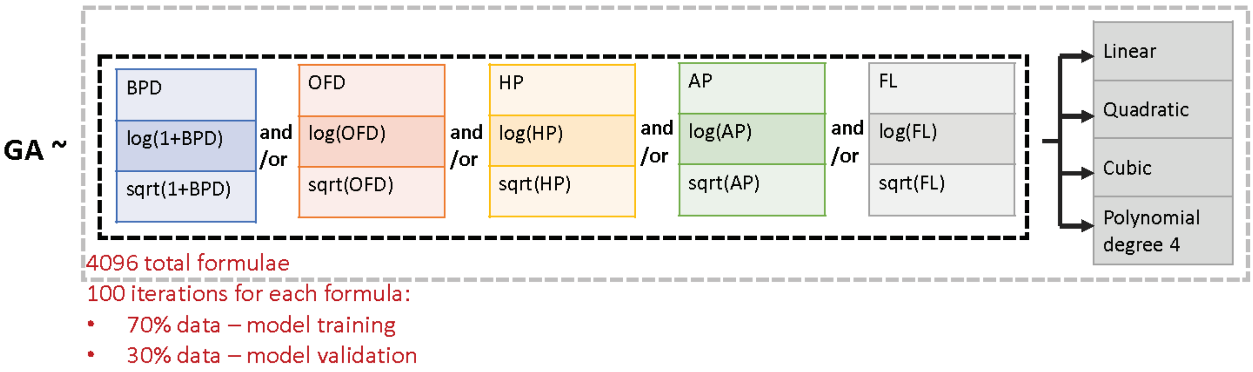


### Figure S3

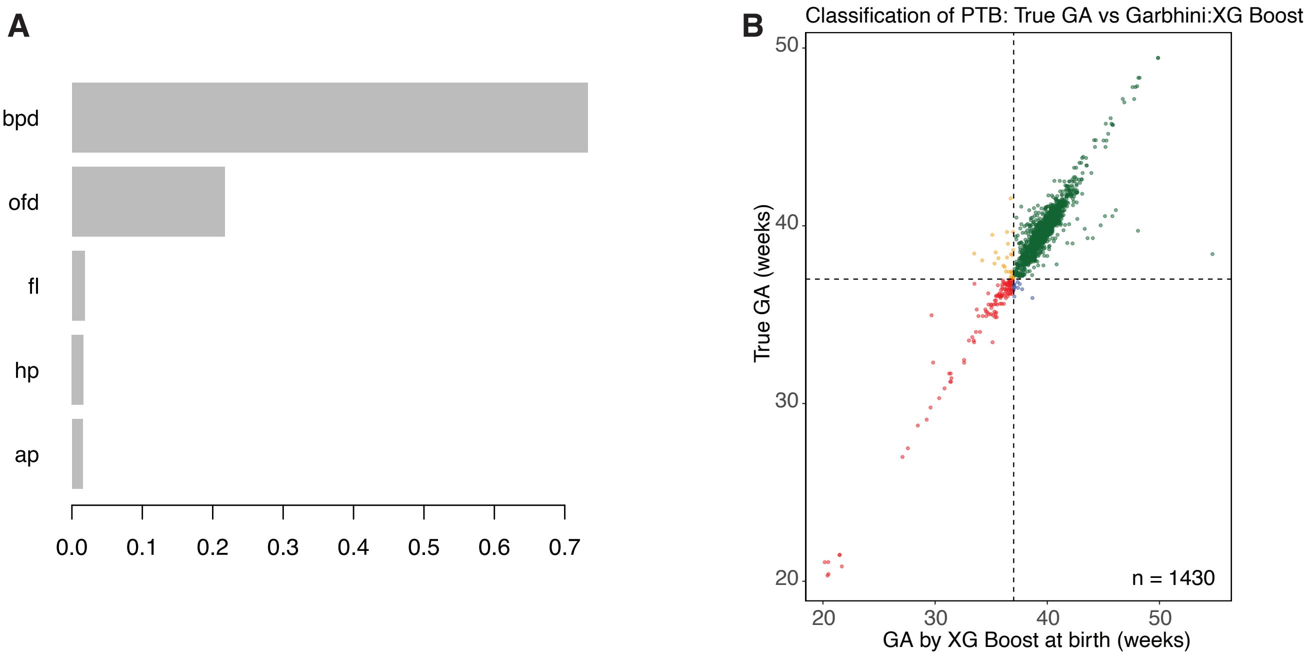

**Figure S4**

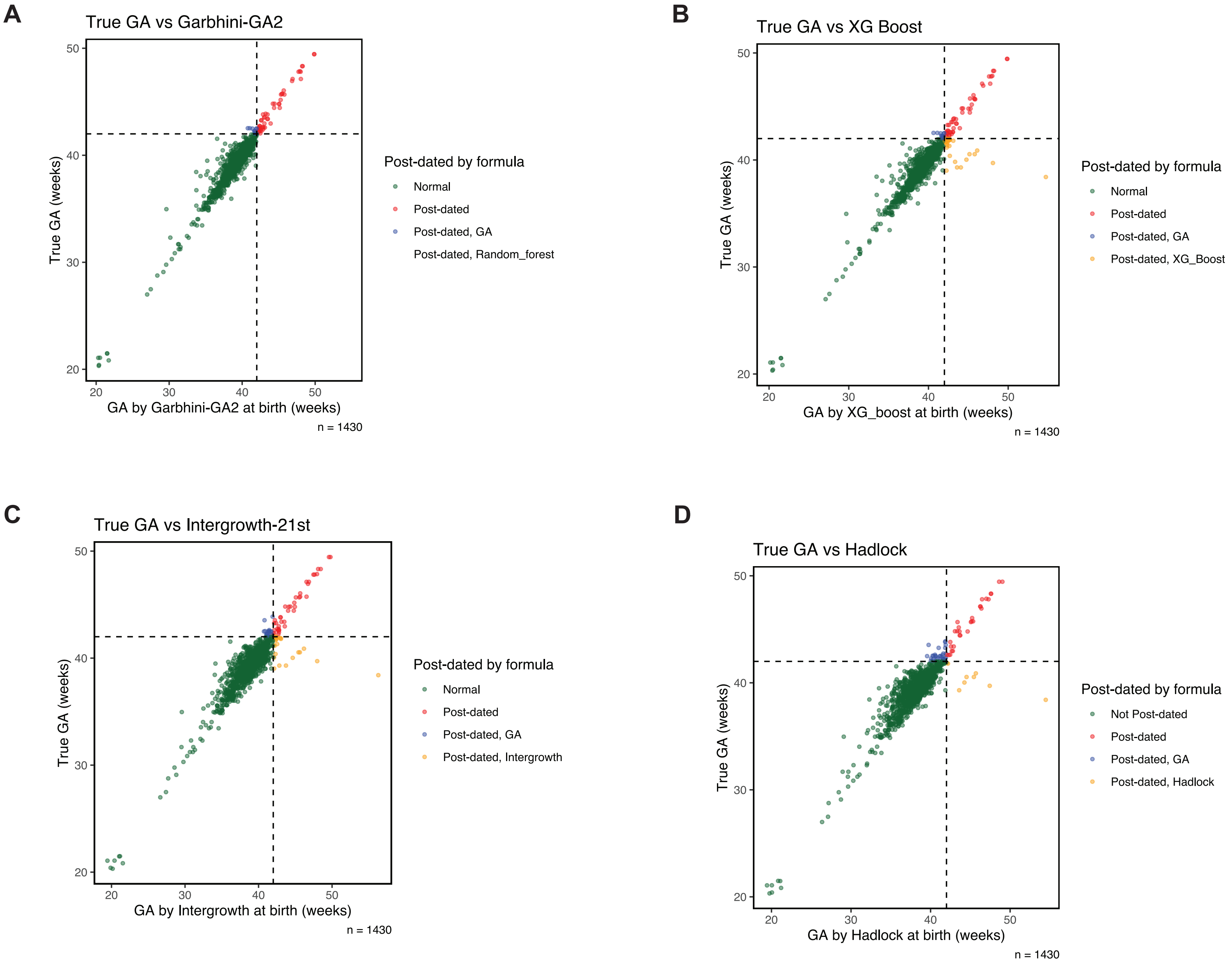
A
True GA vs Garbhini-GA2
True GA (weeks)
GA by Garbhini-GA2 at birth (weeks)
n = 1430
Post-dated by formula
Normal
Post-dated
Post-dated, GA
Post-dated, Random_forest
B
True GA vs XG Boost
True GA (weeks)
GA by XG_boost at birth (weeks)
n = 1430
Post-dated by formula
Normal
Post-dated
Post-dated, GA
Post-dated, XG_Boost
C
True GA vs Intergrowth-21st
True GA (weeks)
GA by Intergrowth at birth (weeks)
n = 1430
Post-dated by formula
Normal
Post-dated
Post-dated, GA
Post-dated, Intergrowth
D
True GA vs Hadlock
True GA (weeks)
GA by Hadlock at birth (weeks)
n = 1430
Post-dated by formula
Not Post-dated
Post-dated
Post-dated, GA
Post-dated, Hadlock

### Figure S5

A

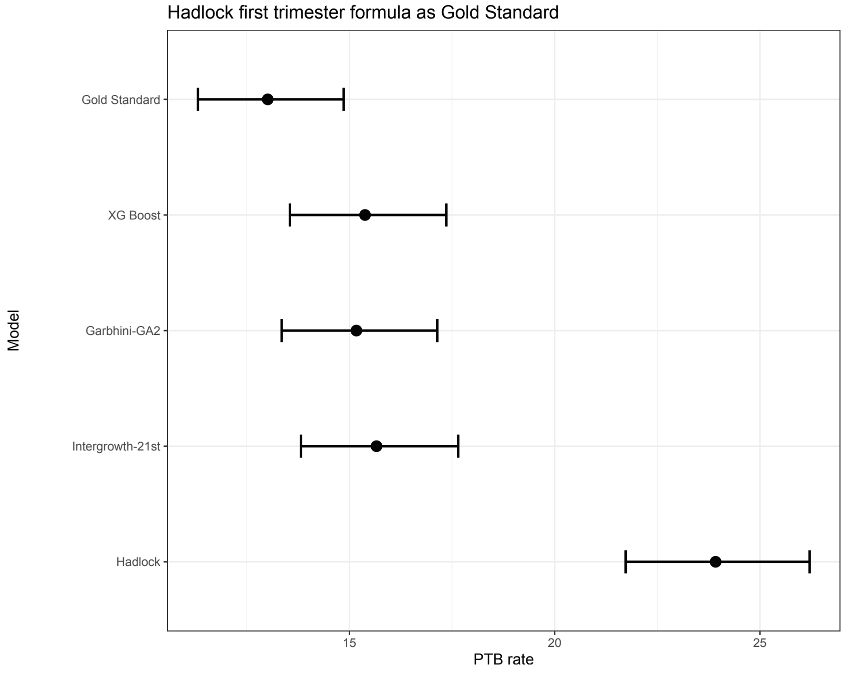

Hadlock first trimester formula as Gold Standard
Gold Standard
XG Boost
Garbhini-GA2
Intergrowth-21st
Hadlock
Model
15
20
25
PTB rate


B

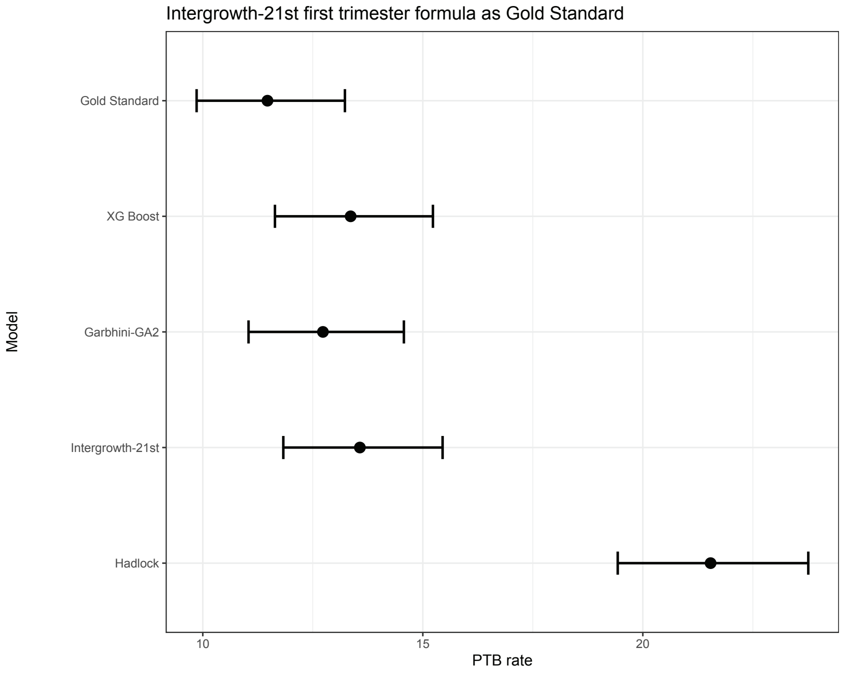

Intergrowth-21st first trimester formula as Gold Standard
Gold Standard
XG Boost
Garbhini-GA2
Intergrowth-21st
Hadlock
Model
10
15
20
PTB rate

**Figure S6**

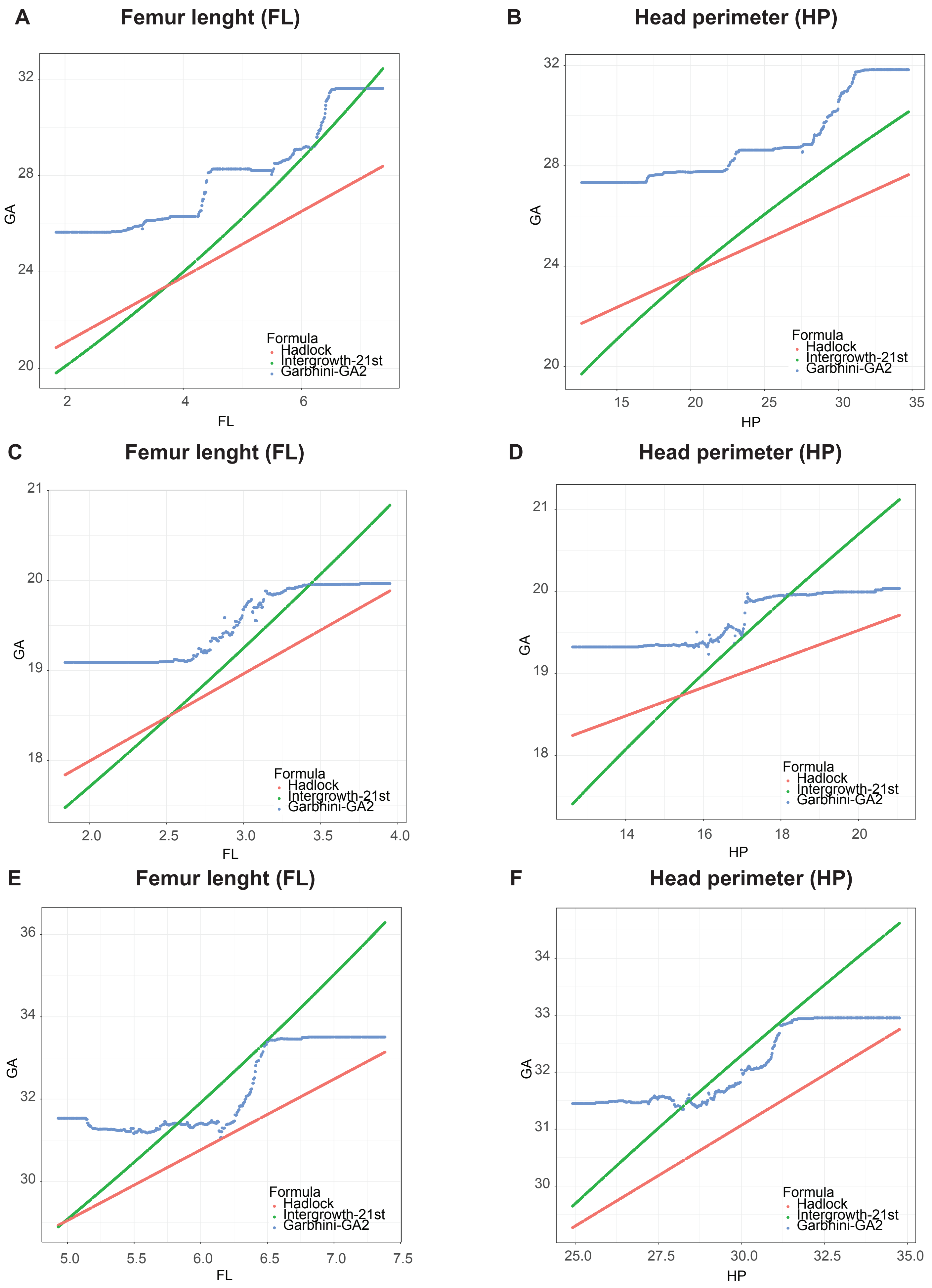
A
Femur lenght (FL)
GA
FL
2
4
6
20
24
28
32
Formula
Hadlock
Intergrowth-21st
Garbhini-GA2
B
Head perimeter (HP)
GA
HP
15
20
25
30
35
20
24
28
32
Formula
Hadlock
Intergrowth-21st
Garbhini-GA2
C
Femur lenght (FL)
GA
FL
2.0
2.5
3.0
3.5
4.0
18
19
20
21
Formula
Hadlock
Intergrowth-21st
Garbhini-GA2
D
Head perimeter (HP)
GA
HP
14
16
18
20
18
19
20
21
Formula
Hadlock
Intergrowth-21st
Garbhini-GA2
E
Femur lenght (FL)
GA
FL
5.0
5.5
6.0
6.5
7.0
7.5
30
32
34
36
Formula
Hadlock
Intergrowth-21st
Garbhini-GA2
F
Head perimeter (HP)
GA
HP
25.0
27.5
30.0
32.5
35.0
30
31
32
33
34
Formula
Hadlock
Intergrowth-21st
Garbhini-GA2

**Supplementary Tables**

**Table S1** List of 21 candidate features, including the five main USG variables (BPD, OFD, HP, AP & FL) used to implement feature selection.

| S No. | Feature list | Description of feature |
|---|---|---|
| 1 | bpd | Biparietal diameter |
| 2 | ofd | Occipitofrontal diameter |
| 3 | hp | Head perimeter |
| 4 | ap | Abdominal perimeter |
| 5 | fl | Femur length |
| 6 | sfh | Symphysio fundal height |
| 7 | part_edu_yrs | Education of husband in years |
| 8 | fmly_inc | Family income |
| 9 | preg_num | Position of current pregnancy with respect to total pregnancies |
| 10 | hemglo | Haemoglobin count |
| 11 | abgircm | Abdominal Girth |
| 12 | anc_cur_wt | Weight of mother |
| 13 | hght | Height of mother |
| 14 | bmi | Body mass index |
| 15 | derived_parity | Parity: number of times a woman has been pregnant |
| 16 | fuel | Indicative of type of fuel used |
| 17 | cl_qc_as1 | Cervical length measured in the second trimester |
| 18 | bp_dia_fup1 | Blood pressure (diastolic) |
| 19 | left_ute_art_pl_as1 | Arterial pressure |
| 20 | contr_bcp | Use of contraceptives prior to current pregnancy |
| 21 | smok_prs | Exposure to second-hand smoke |

**Table S2** Performance characteristics of the GA estimation models as compared to the Gold Standard GA on TEST set. Extended table with comparison of other models

| **Model** | **RMSE** |
|---|---|
| Hadlock | 1.57 [1.52, 1.60] |
| INTERGROWTH-21st | 1.16 [1.08, 1.23] |
| Garbhini (Polynomial regression) | 1.07 [0.99, 1.13] |
| Garbhini-GA2 | 0.89 [0.80, 0.97] |
| Garbhini-XG_Boost | 0.9 [0.82, 0.98] |

**Table S3** Bland Altman analysis and preterm birth agreement on TEST set. Each value in the bottom quadrant consists of a pairwise mean difference between the formulae with each other and with respect to the Gold Standard GA. The value in brackets represents the limits of agreement. Each value in the top quadrant is the agreement between the gold standard and three formulae for preterm labelling computed using the Jaccard similarity coefficient. Extended table with comparison of other models

| | Gold Standard | Garbhini XG Boost | Garbhini-GA2 | Hadlock | INTERGROWTH-21st |
|---|---|---|---|---|---|
| Gold Standard | | 76.074 | 78.750 | 41.486 | 60.280 |
| Garbhini XG Boost | 0.035( -1.731, 1.8) | | 91.772 | 47.205 | 71.429 |
| Garbhini-GA2 | 0.027( -1.718, 1.772) | 0.007(-0.279, 0.293) | | 46.894 | 72.596 |
| Hadlock | 1.089( -1.125, 3.302) | 1.054( -0.242, 2.35) | 1.061( -0.251, 2.374) | | 64.596 |
| INTERGROWTH-21st | 0.447( -1.66, 2.554) | 0.412( -0.7, 1.525) | 0.42( -0.702, 1.541) | -0.642( -1.599, 0.315) | |

**Table S4** Percentage of preterm labelled by each model. Each value corresponds to the percentage of observations with the 95% confidence interval labelled as preterm for each model in the TEST set ($N_o$ = 1379).

| **Model** | **% preterm** | **95% CI.** |
|---|---|---|
| Gold Standard GA | 9.4 | [8.0, 11.1] |
| Hadlock | 22.5 | [20.4,24.8] |
| INTERGROWTH-21st | 14.5 | [12.8,16.5] |
| Garbhini-GA2 | 10.6 | [9.0,12.3] |
| Garbhini XG Boost | 10.6 | [9.1,12.3] |

**Table S5** Percentage of post-term labelled by each model. Each value corresponds to the percentage of observations with the 95% confidence interval labelled as post-term for each model in the TEST set ($N_o$ = 1,379).

| Model | % post-term | 95% CI. |
|---|---|---|
| Gold Standard GA | 4.3 | [3.3, 5.4] |
| Hadlock | 2.7 | [1.9, 3.7] |
| INTERGROWTH-21st | 4.5 | [3.5, 5.7] |
| Garbhini XG Boost | 5.7 | [4.6,7.1] |
| Garbhini-GA2 | 5.8 | [4.6, 7.1] |

**Table S6** Post-term birth agreement on TEST set ($N_o$ = 1,379). Each value in the bottom quadrant is the agreement between the gold standard and three formulae for post-term labelling computed using the Jaccard similarity coefficient.

| | Hadlock | Intergrowth | Garbhini Random Forest | Garbhini XG Boost | Gold-Standard |
|---|---|---|---|---|---|
| **Hadlock** | | | | | |
| **Intergrowth** | 60.938 | | | | |
| **Garbhini Random Forest** | 46.988 | 68.966 | | | |
| **Garbhini XG Boost** | 47.561 | 69.767 | 94.118 | | |
| **Gold-Standard** | 42.857 | 50.602 | 54.839 | 50.526 | |

**Table S7** Comparisons between different dating models when alternate gold standards. RMSE is the root mean squared error of GA estimated by each model compared with respective Gold standard GA in weeks on TEST set ($N_o$ = 1,379).

| | **Hadlock first trimester - Gold Standard** | **INTERGROWTH-21$^{st}$ first trimester – Gold Standard** |
|---|---|---|
| **Model** | RMSE | RMSE |
| **Hadlock** | 1.2419236 | 1.2070661 |
| **INTERGROWTH – 21$^{st}$** | 0.9259802 | 0.9105423 |
| **Garbhini (Brute Force)** | 0.9402165 | 0.9272075 |
| **Garbhini-GA2** | 0.7681168 | 0.75268 |
| **Garbhini-XG_Boost** | 0.7696569 | 0.7586046 |

**Table S8** Comparisons between continuous and categorical variables showing 1-week deviation in predictions for Garbhini-GA2 and Hadlock's model. For continuous and categorical features, Mean (SD) followed by a Welch's two-sample t-test and proportion (%) followed by a Fisher's exact test were calculated, respectively.

| Number | Characteristic | **beyond_1week**, N = 32[1] | **within_1week**, N = 653[1] | **p-value**[2] |
|---|---|---|---|---|
| 1 | bpd | 3.93 (0.19) | 4.39 (0.26) | <0.001 |
| 2 | ofd | 5.42 (0.27) | 5.75 (0.34) | <0.001 |
| 3 | hp | 14.97 (0.77) | 16.26 (0.92) | <0.001 |
| 4 | ap | 12.32 (0.80) | 13.58 (0.88) | <0.001 |
| 5 | fl | 2.65 (0.25) | 2.91 (0.22) | <0.001 |
| 6 | sfh | 15.4 (3.6) | 15.9 (3.1) | 0.4 |
| | Unknown | 2 | 32 | |
| 7 | bmi | 20.9 (2.9) | 21.1 (3.9) | 0.6 |
| | Unknown | 1 | 9 | |
| 8 | abgircm | 74 (8) | 75 (9) | 0.4 |
| | Unknown | 2 | 13 | |
| 9 | hght | 151.2 (6.4) | 152.6 (6.4) | 0.3 |
| 10 | bp_dia_fup1 | 69 (7) | 68 (8) | 0.4 |
| | Unknown | 0 | 24 | |
| 11 | bp_sys_fup1 | 110 (7) | 108 (9) | 0.4 |
| | Unknown | 0 | 34 | |
| 12 | fmly_mem | 4 (2) | 4 (3) | >0.9 |
| 13 | hemglo | 11.27 (12.44) | 12.41 (14.76) | 0.6 |
| | Unknown | 2 | 4 | |
| 14 | derived_parity | | | 0.7 |
| | 0 | 14 / 32 (44%) | 336 / 653 (51%) | |
| | 1 | 11 / 32 (34%) | 209 / 653 (32%) | |
| | 2 | 6 / 32 (19%) | 82 / 653 (13%) | |
| | 3 | 1 / 32 (3.1%) | 22 / 653 (3.4%) | |
| | 4 | 0 / 32 (0%) | 4 / 653 (0.6%) | |
| 15 | fmly_typ | | | 0.2 |
| | 11 | 17 / 32 (53%) | 352 / 653 (54%) | |
| | 12 | 5 / 32 (16%) | 43 / 653 (6.6%) | |
| | 13 | 1 / 32 (3.1%) | 20 / 653 (3.1%) | |
| | 14 | 9 / 32 (28%) | 238 / 653 (36%) | |
| 16 | rlgn | | | 0.3 |
| | 11 | 27 / 32 (84%) | 597 / 653 (91%) | |

| | | | | |
|---|---|---|---|---|
| | 12 | 4 / 32 (12%) | 44 / 653 (6.7%) | |
| | 13 | 0 / 32 (0%) | 4 / 653 (0.6%) | |
| | 14 | 1 / 32 (3.1%) | 5 / 653 (0.8%) | |
| | 15 | 0 / 32 (0%) | 2 / 653 (0.3%) | |
| | 17 | 0 / 32 (0%) | 1 / 653 (0.2%) | |
| 17 | part_occ | | | 0.06 |
| | 11 | 26 / 32 (81%) | 605 / 653 (93%) | |
| | 12 | 5 / 32 (16%) | 25 / 653 (3.8%) | |
| | 13 | 0 / 32 (0%) | 10 / 653 (1.5%) | |
| | 14 | 1 / 32 (3.1%) | 6 / 653 (0.9%) | |
| | 15 | 0 / 32 (0%) | 1 / 653 (0.2%) | |
| | 16 | 0 / 32 (0%) | 1 / 653 (0.2%) | |
| | 17 | 0 / 32 (0%) | 5 / 653 (0.8%) | |
| 18 | drnk_wtr | | | 0.5 |
| | 11 | 14 / 32 (44%) | 288 / 653 (44%) | |
| | 12 | 5 / 32 (16%) | 97 / 653 (15%) | |
| | 13 | 9 / 32 (28%) | 157 / 653 (24%) | |
| | 14 | 0 / 32 (0%) | 1 / 653 (0.2%) | |
| | 16 | 0 / 32 (0%) | 4 / 653 (0.6%) | |
| | 19 | 3 / 32 (9.4%) | 102 / 653 (16%) | |
| | 21 | 1 / 32 (3.1%) | 3 / 653 (0.5%) | |
| | 22 | 0 / 32 (0%) | 1 / 653 (0.2%) | |
| 19 | derived_ses_mks_2019 | | | 0.4 |
| | 0 | 0 / 31 (0%) | 5 / 650 (0.8%) | |
| | 1 | 10 / 31 (32%) | 122 / 650 (19%) | |
| | 2 | 9 / 31 (29%) | 222 / 650 (34%) | |
| | 3 | 12 / 31 (39%) | 298 / 650 (46%) | |
| | 4 | 0 / 31 (0%) | 3 / 650 (0.5%) | |
| | Unknown | 1 | 3 | |
| 20 | tob_chew | | | 0.3 |
| | 11 | 31 / 32 (97%) | 647 / 652 (99%) | |
| | 12 | 0 / 32 (0%) | 1 / 652 (0.2%) | |
| | 13 | 0 / 32 (0%) | 1 / 652 (0.2%) | |
| | 14 | 1 / 32 (3.1%) | 3 / 652 (0.5%) | |
| | Unknown | 0 | 1 | |
